# Association between long working hours and psychological distress: The effect of sick leave criteria in the workplace during the COVID-19 pandemic

**DOI:** 10.1101/2021.11.07.21266036

**Authors:** Ayako Hino, Akiomi Inoue, Kosuke Mafune, Mayumi Tsuji, Seiichiro Tateishi, Akira Ogami, Tomohisa Nagata, Keiji Muramatsu, Yoshihisa Fujino, for the CORoNaWork Project

**Affiliations:** Department of Mental Health, Institute of Industrial Ecological Sciences, University of Occupational and Environmental Health, Japan; Institutional Research Center, University of Occupational and Environmental Health, Japan; Department of Environmental Health, School of Medicine, University of Occupational and Environmental Health, Japan; Department of Occupational Medicine, School of Medicine, University of Occupational and Environmental Health, Japan; Department of Work Systems and Health, Institute of Industrial Ecological Sciences, University of Occupational and Environmental Health, Japan; Department of Occupational Health Practice and Management, Institute of Industrial Ecological Sciences, University of Occupational and Environmental Health, Japan; Department of Preventive Medicine and Community Health, School of Medicine, University of Occupational and Environmental Health, Japan; Department of Environmental Epidemiology, Institute of Industrial Ecological Sciences, University of Occupational and Environmental Health, Japan

**Keywords:** Long working hours, Psychological distress, COVID-19, Cross-sectional studies, Japan

## Abstract

**Objective:** This study investigated the effect of sick leave criteria on the association between long working hours and psychological distress.

**Methods:** We conducted a cross-sectional survey in December 2020, and 27,032 workers completed the questionnaire. First, after testing the interaction effect of overtime work hours and sick leave criteria on psychological distress, we conducted stratified analyses using sick leave criteria.

**Results:** A significant interaction effect was found. When we conducted stratified analyses, the odds ratios increased with longer working hours, both with and without sick leave criteria groups; however, the risk was greater in the without sick leave criteria group, compared with the criteria group.

**Conclusion:** We revealed that working without sick leave criteria could strengthen the association between long working hours and psychological distress during the COVID-19 pandemic.

**Clinical significance:** Workers working without sick leave criteria had a significantly higher risk of psychological distress due to long working hours than those who worked with the criteria. Our findings contribute to preventing the deterioration of mental health during the COVID-19 pandemic.

## Introduction

The coronavirus disease 2019 (COVID-19), a respiratory disease caused by SARS-COV-2, has resulted in a global pandemic. In Japan, the first case of infection was reported in early 2020; the infection spread rapidly, thereafter. The first epidemic wave emerged in April 2020, the second in July 2020, and the third in December 2020; moreover, while ongoing waves continue up until today. The Japanese government implemented community-based measures, similar to those applied globally, to control the pandemic, including quarantining, encouraging people’s prevention practices, restricting travel and events, and promoting social distancing^1^. However, these infection control measures should be implemented by not only individuals but also companies. Therefore, several workplace measures are implemented to control COVID-19 infections worldwide.

Several previous studies in Japan have reported the association between workplace infection measures and mental health. A cross-sectional study reported that several workplace measures result in better psychological distress and job performance^2^. Another cross-sectional study revealed that companies with few measures had significantly worse mental health compared with those with sufficient measures^3^. These previous studies included multiple workplace infection measures such as preventive measures taken including those implemented by individuals (e.g., hand washing, wearing masks), preventive measures to reduce the risk of infection at the workplace (e.g., refrain from business trips, restricting outside visitors), and criteria and procedures for waiting at home and clinical contact (e.g., request to refrain from going to work when ill, report request for fever). Among others, Sasaki et al^2^ reported that “criteria and procedures for waiting at home and clinical contact” significantly increased job performance. Another study reported a similar tendency, which showed that “requesting employees not come to work when they are not feeling well” was highly associated with decreased psychological distress among workplace infection control measures^3^. Among other workplace infection control measures, setting the sick leave criteria (i.e., criteria for deciding whether or not employees come to work when they are unwell) may be particularly important for mental health.

On the other hand, long working hours are considered a problem in Japanese workplaces. “Karoshi” (death from overwork) and “Karojisatsu” (suicide induced by overwork) have been prevalent social problems in Japan for the past several decades. The association between long working hours and physical health has been reported in some previous studies^4^. Moreover, similar evidence was reported for mental health in recent years^5^.

While numerous studies during the COVID-19 pandemic have revealed the association between long working hours and mental health, a majority of such studies have been conducted on healthcare workers. For example, a cross-sectional study in a Japanese healthcare institution showed that increasing working hours might be a risk factor for depression among nurses^6^. Further, a prior cross-sectional study conducted in foreign countries (i.e., not including Japan) found that job strain, including increased working hours, was the most significant psychosocial stressor among German healthcare workers^7^. However, during the COVID-19 pandemic, along with healthcare workers, other general workers may also be exposed to overworking. The International Labour Organization (ILO) guidelines^8^ lists long working hours as an important psychosocial risk during the COVID-19 pandemic, indicating that the psychological effects of long working hours are an important issue for non-healthcare workers.

Furthermore, during the COVID-19 pandemic, long working hours may increase the possibility of infection, resulting in a strong association between long working hours and psychological distress in workplaces with inadequate infection control measures. However, the effect of workplace infection control measures on the association between long working hours and psychological distress has not yet been revealed. In this study, we particularly focused on sick leave criteria and investigated the effect of the criteria on the association between long working hours and psychological distress. We hypothesized that the association between long working hours and psychological distress would be stronger for workers in workplaces without sick leave criteria than those who working with such criteria.

## Methods

### Participants

We conducted a cross-sectional survey from December 22–26, 2020, among participants who had previously registered with a Japanese web survey company. At the time of this survey, the third wave of the COVID-19 pandemic had occurred in Japan, and the number of infected persons was increasing rapidly. Details of the protocol for this study have been previously reported^9^. The participants were selected by allocating an equal distribution of numbers by gender, occupation, and residential region, and 33,302 were eligible to respond. Participants with exceptionally short response times (≤ 6 minutes), exceptionally low body weight (< 30 kg), exceptionally short height (< 140 cm), and those who provided inconsistent responses to comparable questions (e.g., inconsistent responses to questions on marital status and area of residence), as well as incorrect responses to a question (i.e., those who gave fraudulent responses; select the third largest number from among the following five numbers) were excluded. After excluding these invalid responses, the total number of respondents included in the final analysis was 27,032. Participants’ demographic and occupational characteristics are presented in Table 1.

**Table 1.** Participants’ demographic and occupational characteristics, overtime work hours, and psychological distress by sick leave criteria (n = 27,036)

| | With sick leave criteria<br>(n = 20,230) | | Without sick leave criteria<br>(n = 6,806) | | Cronbach's $\alpha$ |
| --- | --- | --- | --- | --- | --- |
|  | Mean (SD) | n (%) | Mean (SD) | n (%) |  |
| Gender |  |  |  |  |  |
| Men |  | 10,025 (49.6) |  | 3,789 (55.7) |  |
| Women |  | 10,205 (50.4) |  | 3,017 (44.3) |  |
| Age | 46.7 (10.6) |  | 47.9 (10.3) |  |  |
| Marital status |  |  |  |  |  |
| Married |  | 11,625 (57.5) |  | 3,406 (50.0) |  |
| Divorce, widowed |  | 2,035 (10.1) |  | 808 (11.9) |  |
| Unmarried |  | 6,572 (32.5) |  | 2,592 (38.1) |  |
| Residence status |  |  |  |  |  |
| Living with family |  | 15,989 (79.0) |  | 5,240 (77.0) |  |
| Living alone |  | 4,241 (21.0) |  | 1,566 (23.0) |  |
| Educational background |  |  |  |  |  |
| Junior high school |  | 214 (1.1) |  | 154 (2.3) |  |
| High school |  | 4,805 (23.8) |  | 2,148 (31.6) |  |
| Vocational school |  | 2,643 (13.1) |  | 1,030 (15.1) |  |
| Junior college, technical college |  | 2,188 (10.8) |  | 683 (10.0) |  |
| University |  | 9,187 (45.4) |  | 2,534 (37.2) |  |
| Graduate school |  | 1,193 (5.9) |  | 257 (3.8) |  |
| Present illness |  |  |  |  |  |
| None |  | 13,057 (64.5) |  | 4,469 (65.7) |  |
| Any |  | 7,173 (35.5) |  | 2,337 (34.3) |  |
| Residence area |  |  |  |  |  |
| High infection rate areas |  | 8,375 (41.4) |  | 2,827 (41.5) |  |
| Low infection rate areas |  | 11,855 (58.6) |  | 3,979 (58.5) |  |
| Job type |  |  |  |  |  |
| Mainly desk work |  | 10,303 (50.9) |  | 3,165 (46.5) |  |
| Jobs mainly involving interpersonal communication |  | 5,267 (26.0) |  | 1,660 (24.4) |  |
| Mainly labor |  | 4,660 (23.0) |  | 1,981 (29.1) |  |
| Frequency of remote working |  |  |  |  |  |
| $\geq 4$ times/week | | 1,702 (8.4) | | 1,088 (16.0) | |
| $\geq 2$ times/week | | 1,236 (6.1) | | 241 (3.5) | |
| $\geq 1$ time/week | | 745 (3.7) | | 133 (2.0) | |
| $\geq 1$ time/month | | 530 (2.6) | | 85 (1.2) | |
| Almost none |  | 16,017 (79.2) |  | 5,259 (77.3) |  |
| Psychosocial work characteristics (JCQ)<sup>†</sup> |  |  |  |  |  |
| Job Control | 63.5 (11.3) |  | 63.0 (10.3) |  | 0.74 |
| Supervisor support | 10.4 (2.8) |  | 9.0 (3.3) |  | 0.94 |
| Coworker support | 10.8 (2.4) |  | 9.7 (3.0) |  | 0.90 |
| Overtime work hours (daily average) |  |  |  |  |  |
| Almost none |  | 8,945 (44.2) |  | 3,898 (57.3) |  |
| < 2 hours |  | 9,417 (46.5) |  | 2,323 (34.1) |  |
| $\geq 2$ hours | | 1,868 (9.2) | | 585 (8.6) | |
| Psychological distress (K6) | 4.4 (5.2) |  | 5.5 (6.0) |  | 0.93 |
| Number of cases<sup>‡</sup> |  | 7,678 (38.0) |  | 3,139 (46.1) |  |

<sup>†</sup> JCQ, Job Content Questionnaire.<sup>‡</sup> Psychological distress was defined as having a score of five or more on the K6 scale.

The purpose and procedures of the study were explained to the participants, and informed consent was obtained through an online website. The Ethics Committee of Medical Research, University of Occupational and Environmental Health, Japan, reviewed and approved the study procedures (approval number: R2-079).

### Measures

#### 1) Overtime work hours

We obtained information on usual daily overtime work hours using a self-administered questionnaire. We explored overtime using the question, “How many hours of overtime do you work per day?” Following the classification standard used in a previous systematic review of overtime work hours^5^, overtime work hours per day were classified into three groups: almost none, < 2 hours, and ≥ 2 hours, which is equivalent to less than 40 hours, < 50 hours, and ≥ 50 hours of working hours per week, respectively.

#### 2) Sick leave criteria

A single-item scale was used to investigate the sick leave criteria using the question, “Does your workplace request employees to refrain from coming in to work when they are not feeling well?” in the questionnaire. We defined those responding with “yes” as “with sick leave criteria” and those answering “no” as “without sick leave criteria.”

#### 3) Psychological distress

In this survey, psychological distress was measured using the K6 scale^10, 11^. This scale has been widely used in Japan as well as in foreign countries to measure the symptoms of psychological distress in the past 30 days. The K6 scale comprises a six-item scale measuring the extent of psychological distress in the past 30 days with a five-point response option ranging from 0 = *none of the time* to 4 = *all of the time* (response range, 0–24). In this sample, Cronbach’s α coefficient was 0.93. Participants were dichotomized into those with psychological distress (total K6 score of five or more) and those without (0–4 score) using a cutoff point recommended for the Japanese population^12^.

#### 4) Other covariates

Other covariates included demographic characteristics (i.e., gender, age, marital status, residence status, educational background, present illness, and residence area) and occupational characteristics (i.e., job type, frequency of remote working, and psychosocial work characteristics). Marital status was classified into three groups: married, divorced or widowed, and unmarried. Residence status was classified into two groups: living with family and living alone. Educational background was classified into six groups: junior high school, high school, vocational school, junior college or technical college, university, and graduate school. Present illness was classified into two groups: none and any. Residence areas were classified into two groups: high infection rate areas (i.e., prefectures with a declaration of a state of emergency by the Japanese Government due to COVID-19^13^) and low infection rate areas (i.e., prefectures without a declaration of a state of emergency by the Japanese Government due to COVID-19). Job type was classified into three groups: mainly desk work, jobs mainly involving interpersonal communication, and mainly labor. The frequency of remote working was classified into five groups: ≥ 4 times/week, ≥ 2 times/week, ≥ 1 time/week, ≥ 1 time/month, and almost none. Psychosocial work characteristics, including job control, supervisor support, and coworker support, were assessed using the Japanese version of the Job Content Questionnaire (JCQ)^14, 15^. The JCQ includes a nine-item job control scale (response range 24–96, Cronbach’s alpha in the present sample = 0.74), a four-item coworker support scale (response range 4–16, Cronbach’s alpha in the present sample = 0.94), and a four-item coworker support scale (response range 4–16, Cronbach’s alpha in the present sample = 0.90).

### Statistical analysis

We conducted multiple logistic regression analyses to estimate the prevalence of odds ratios (ORs) and their 95 % confidence intervals (CIs) of psychological distress (defined as having a score of five or more on the K6 scale). In the analyses, we first tested the interaction effect of overtime work hours and sick leave criteria on psychological distress. When we observed a significant interaction effect, we conducted stratified analyses using the sick leave criteria. In a series of analyses, we first calculated the crude ORs (i.e., without any adjustment) (Model 1). Then, we adjusted for demographic characteristics (i.e., gender, age, marital status, residence status, educational background, present illness, and residence area) (Model 2) and additionally adjusted for occupational characteristics (i.e., job type, frequency of remote working, and psychosocial work characteristics) (Model 3). The level of significance was set at *p* < 0.05. Statistical analyses were performed using IBM Statistical Package for the Social Sciences (SPSS) Statistics version 22 (SPSS Inc., Chicago, IL, USA).

## Results

Of all participants, 20,230 (74.8 %) worked with sick leave criteria and 6,806 (25.2%) worked without sick leave criteria. Compared with the participants with sick leave criteria, those without sick leave criteria were more likely to have no overtime work and had higher K6 scores. Furthermore, participants without sick leave criteria were more likely to be male, unmarried, living alone, mainly engaged in labor, and have a lower educational background than those with sick leave criteria.

The interaction effects of overtime work hours and sick leave criteria on psychological distress were significant in any models (*p* for interaction < 0.001) (data not shown). When we conducted stratified analyses by the sick leave criteria, in the “with sick leave criteria” group, the < 2 hours subgroup (ORs = 1.31–1.32; 95 % CIs: 1.24–1.41), and ≥ 2 hours subgroup (ORs = 1.60–1.74; 95 % CIs: 1.45–1.94) had significantly higher ORs of psychological distress compared with the almost no overtime subgroup in all models (Table 2). A similar tendency was observed in the without sick leave criteria group; however, the ORs were greater in the without sick leave criteria group than the with sick leave criteria group.

**Table 2.** Association between overtime work hours and psychological distress by sick leave criteria: multiple logistic regression analysis

| Overtime work hours | *n* | No. of cases (%) | Odds ratio (95 % confidence interval) |  |  |
| --- | --- | --- | --- | --- | --- |
|  |  |  | Model 1 † | Model 2 ‡ | Model 3 § |
| With sick leave criteria |  |  |  |  |  |
| Almost none | 8,945 | 3,038 (34.0) | 1.00 | 1.00 | 1.00 |
| < 2 hours | 9,417 | 3,796 (40.3) | 1.31 (1.24–1.39) | 1.32 (1.24–1.40) | 1.32 (1.24–1.41) |
| ≥ 2hours | 1,868 | 844 (45.2) | 1.60 (1.45–1.77) | 1.74 (1.56–1.94) | 1.73 (1.55–1.93) |
|  |  |  | *p* < 0.001 | *p* < 0.001 | *p* < 0.001 |
| Without sick leave criteria |  |  |  |  |  |
| Almost none | 3,898 | 1,547 (39.7) | 1.00 | 1.00 | 1.00 |
| < 2 hours | 2,323 | 1,266 (54.5) | 1.82 (1.64–2.02) | 1.75 (1.57–1.95) | 1.77 (1.58–1.98) |
| ≥ 2hours | 585 | 326 (55.7) | 1.91 (1.61–2.28) | 1.95 (1.63–2.35) | 1.95 (1.62–2.36) |
|  |  |  | *p* < 0.001 | *p* < 0.001 | *p* < 0.001 |

† Crude (i.e., without any adjustment).
‡ Adjusted for demographic characteristics (i.e., gender, age, marital status, residence status, educational background, present illness, and residence area).
§ Additionally adjusted for occupational characteristics (i.e., job type, frequency of remote working, and psychosocial work characteristics).
Psychological distress was defined as having a score of five or more on the K6 scale.
*p* for interaction (overtime work hours \* sick leave criteria) < 0.001 in any models.

## Discussion

This study demonstrated a significant interaction effect of overtime work hours and sick leave criteria on psychological distress. Workers without sick leave criteria had a significantly higher risk of psychological distress due to long working hours than those who worked with the criteria. As mentioned earlier, previous studies reported that long working hours resulted in deteriorating mental health during the COVID-19 pandemic^6, 7^, and the setting of sick leave criteria would be particularly important for mental health among other COVID-19 infection control measures^2, 3^. Our study extends prior findings by revealing that the absence of sick leave criteria in the workplace could result in a stronger association between long working hours and psychological distress. Workers working long hours experience poor quality and quantity of sleep, fatigue, and disruption of family and social activities, which results in poor mental health^16^. Long working hours, during a pandemic, may have an additional impact on poor mental health due to the increasing possibility of infection because of the prolonged time spent in the workplace. During the influenza A (H1N1) pdm09 outbreak in 2009, a workplace policy to allow sick employees to take leave was associated with a lower likelihood of overall workplace infections^17, 18^. Thus, working with sick leave criteria during a pandemic may have had a better impact on mental health by decreasing the anxiety of contracting an infection at work. Therefore, in the absence of sick leave criteria, the association between long working hours and mental health may become stronger during the COVID-19 pandemic.

However, it should be noted that setting sick leave criteria does not completely buffer psychological distress owing to long working hours. The overtime group (i.e., < 2 hours and ≥ 2 hours subgroups) had higher ORs, compared with the group with no overtime, even among workers with the criteria. Thus, workplaces should not only set criteria but also promote the reduction of long working hours during the COVID-19 pandemic.

Some limitations of this study should be considered. First, this was a cross-sectional study; therefore, causality is unclear. Moreover, those with mental health issues may work longer hours because of reduced work efficiency. However, in general, in companies in Japan, occupational health staff is assigned to workplaces to ensure that those with mental health issues usually do not work long hours. Therefore, while it is unlikely that causality will be reversed, future longitudinal studies are needed. Second, since we assessed overtime work hours using self-administered questionnaires, which may have resulted in a bias. However, a previous study revealed that the correlation between self-reported and company records of working hours was quite high^19^; thus, the effect may not be large. Third, to assess overtime work hours, we examined overtime work hours per day; however, previous studies used the average overtime work hours per month or per week^20, 21^. Thus, the workload evaluation may not be accurate in the case of occupations with a greater change in daily working hours. Moreover, we adjusted for the job type, but we did not survey the details of the occupation. Fourth, we did not survey “whether the workplace compensates sick pay during absence.” Since sick pay allows employees to take time off from work without worry, a lack of sick pay could worsen mental health^8^. As the sick leave criteria measured in this study may include those without sick pay during absence, the amplifying effect of an absence of sick leave criteria found in this study may be underestimated. Thus, future studies should not only investigate the criteria of sick leave from work but also sick pay for absence from work. Fifth, there was a lack of information regarding the possible confounding factors affecting mental health. Workplaces with sick leave criteria are likely to have better workplace factors (e.g., good benefits and good job descriptions). Although we adjusted for job type, frequency of remote working, and psychosocial work characteristics, the effects may not be fully eliminated. Future research should also investigate additional workplace factors to eliminate their influence.

In conclusion, this study revealed an amplifying effect of without sick leave criteria on the association between long working hours and psychological distress. We revealed that working without sick leave criteria could strengthen the association between long working hours and mental health during the COVID-19 pandemic. The ILO guideline^8^ and guidelines based on the National Institute for Occupational Safety and Health (NIOSH) Total Worker Health (TWH) program^22^ highlight the importance of informing workers about sick leave policies, in addition to preventing long working hours. Our findings support the ILO and NIOSH TWH guidelines and are useful for preventing the deterioration of mental health during a pandemic.

## Data Availability

All data produced in the present study are available upon reasonable request to the authors.

## Acknowledgements

The current members of the CORoNaWork Project, in alphabetical order, are as follows: Dr Yoshihisa Fujino (present chairperson of the study group), Dr Akira Ogami, Dr Arisa Harada, Dr Ayako Hino, Dr Hajime Ando, Dr Hisashi Eguchi, Dr Kazunori Ikegami, Dr Kei Tokutsu, Dr Keiji Muramatsu, Dr Koji Mori, Dr Kosuke Mafune, Dr Kyoko Kitagawa, Dr Masako Nagata, Dr Mayumi Tsuji, Ms Ning Liu, Dr Rie Tanaka, Dr Ryutaro Matsugaki, Dr Seiichiro Tateishi, Dr Shinya Matsuda, Dr Tomohiro Ishimaru, and Dr Tomohisa Nagata. All members are affiliated with the University of Occupational and Environmental Health, Japan

